# Implementing a Negative Pressure Isolation Space within a Skilled Nursing Facility to Control SARS-CoV-2 Transmission

**DOI:** 10.1101/2020.07.04.20143123

**Authors:** Shelly Miller, Debanjan Mukherjee, Joseph Wilson, Nicholas Clements, Cedric Steiner

## Abstract

**Background:** Isolation space must be expanded during pandemics involving airborne transmission. Little to no work has been done to establish optimal design strategies and implementation plans to ease surge capacity and expand isolation capacity over long periods in congregate living facilities. The COVID-19 pandemic has an airborne transmission component and requires isolation, which is difficult to accomplish in skilled nursing facilities.

**Purpose:** In this study we designed, implemented, and validated an isolation space at a skilled nursing facility in Lancaster, PA. The overall goal was to minimize disease transmission between residents and staff within the facility.

**Basic Procedures:** We created an isolation space by modifying an existing HVAC system of the SNF. We measured pressure on-site and performed computational fluid dynamics and Lagrangian particle-based modeling to test containment and possible transmission extent given the isolation space is considered negative rather than individual rooms.

**Main Findings:** Pressure data shows the isolation space maintained an average hourly value of (standard deviation) -2.3 Pa (0.12 Pa) pressure differential between it and the external hallway connected to the rest of the facility. No transmission of SARS-CoV-2 between residents isolated to the space occurred, nor did any transmission to the staff or other residents occur. The isolation space was successfully implemented and, as of writing, continues to be operational through the pandemic.

**Highlights:**

- Negative pressure isolation space is an effective method to meet needed surge capacity during the COVID-19 pandemic and future pandemics
- Planning for how and where to rapidly create a negative pressure isolation space is needed in congregate living areas such as skilled nursing facilities
- This demonstration shows the feasibility of using low-cost and in-house systems to quickly create negative pressure within a skilled nursing facility hallway and to maintain these conditions, minimizing disease transmission between residents and staff

## Background

It was estimated that in the US there were 15,600 nursing homes in 2016 and 1.3 million residents (NCHS 2020). The COVID-19 pandemic has had a disproportionate effect on skilled nursing facilities (SNFs): about 20% of the US virus deaths are linked to SNFs to date. Some states have experienced, and continue to have, significant outbreaks in SNFs (McMichael et al. 2020). An outbreak in Washington demonstrated that much of the transmission of SARS-CoV-2 within the facility was likely due to asymptomatic cases (Aarons et al. 2020). On March 23, it was reported that there were at least 146 nursing homes across 27 states with confirmed Coronavirus cases (CMS 2020); a week later, on March 30, the Centers for Disease Control and Prevention (CDC) had reported more than 400 facilities with infected residents. The CDC recently released guidelines for preparing for COVID-19 in long-term care facilities and nursing homes (CDC 2020a). In this guidance, it is recommended that a cohort space in the facility be identified to care for residents with confirmed or suspected SARS-CoV-2.

The two main routes of transmission of SARS-CoV-2 are contact with contaminated surfaces and inhalation of airborne virus-containing respiratory particles. The virus has been detected by PCR in the air and on surfaces in several healthcare environments (Chia et al. 2020; Ding et al. 2020, Guo et al. 2020; Jiang et al. 2020; Liu et al. 2020; Lednicky et al. 2020). A study from Wuhan reported that SARS-CoV-2 contaminated surfaces including floors and air in an intensive care unit and a general ward of a hospital (Guo et al. 2020). Virus-laden aerosols were concentrated near and downstream from patients at up to 4 m. Lednicky et al. collected positive air samples from more than 2 m away from the nearest patients in a student healthcare center (Lednicky et al. 2020). Outbreaks have also been reported at restaurants, fitness gyms, churches, and festivals (Lu et al. 2020; Nishiura et al. 2020; Pung et al. 2020).

The risk due to airborne transmission necessitates additional controls. Negative pressure isolation spaces are a well-established approach to reduce airborne transmission and guidelines for design and operation have been provided (ASHRAE 2008; Facility Guidelines Institute 2014). Airborne infection isolation rooms are commonly used for obligate airborne infectious disease containment such as tuberculosis. Considering the scale of COVID-19 outbreak, establishing and maintaining effective negative pressure spaces within congregate healthcare facilities to handle surges of patients form a critical piece for healthcare operations and outbreak management. Recent recommendations from the CDC for infection prevention and control for COVID-19 patients in healthcare settings include isolation recommendations and airborne isolation room use for patients undergoing aerosol-generating procedures (CDC 2020b).

Prior work on surge capacity included a demonstration that converted a 30-bed hospital ward to an airborne infection isolation area, in anticipation of a flu pandemic (Miller et al. 2017). This demonstration showed that -30 Pa could be achieved and maintained for 24 hours and the anteroom could be rapidly established; the workers on the ward however had to wear personal protective equipment due to the change in airflow and pressures within the ward. Additional guidelines for creating temporary negative-pressure or other protective environments have been summarized (Rebmann 2005).

To reduce airborne infection risk, the approach from Miller et al. (2017) was applied to a SNF in Lancaster, PA. In this paper, we describe the design and outcomes of establishing this negative pressure isolation space in one ward of the SNF. Systematic modifications were made to existing building HVAC units. These modifications were not resource intensive and were rapidly established. Continuous pressure differential measurements and computational modeling were used to validate the isolation space performance with respect to containment of airborne particles. With a simple anteroom set-up and quick HVAC unit modifications, we were able to maintain continuous negative pressure and contained airflow for a period of 14 days, and the negative pressure area continues to operate.

## Materials and Methods

### Study Area

The study was conducted at a Life Plan Community located on a knoll at an elevation of 146 m in Lancaster County, Pennsylvania. Residential living, personal care, and skilled nursing care units are located within the community. The total campus census at the beginning of the study was 349 residents and 320 staff. The SNF had 114 beds and represented 96 of these residents. One hall was the subject of this study. This hall was on the bottom floor of a wing of a larger building containing administrative offices and the SNF with two floors, each consisting of four wings totaling eight skilled nursing halls. This building adjoined personal care and apartment residential units. The hall selected for conversion into a negative pressure isolation space previously functioned as a short-term rehabilitation area adjacent to a nursing station. Positioned on the facility’s rear, this hall’s advantageous design eased resident relocation and included a series of fire doors separating it from other interior parts of the larger building. The hall is oriented north-south, with external doors on the north end and a nursing station at the south end. Winds predominate from the west. Recorded outdoor wind speeds average 16 kph, but do not typically exceed 48 kph as recorded from the facility’s weather station.

### Baseline Hall Configuration

The hall used in this study consisted of 13 beds within 7 rooms, six double occupancy rooms (32.5-37.2 m^2^) and one single occupancy room (27.0 m^2^). The architectural design of the isolation hall is presented in Figure 1 (left panel). Each room had a single bathroom. The single occupancy room included an anteroom containing a separate sink and medical cabinet. Rooms had a sealed solid glass window facing the hall (100 cm tall × 50 cm wide) and an exterior window (1.85 m tall × 1.35 m wide). Heating and cooling were self-contained in each room with designated units, fans, and air ducts. During normal operations, vents (10.2 cm × 10.2 cm) located in the bathrooms handled exhaust, which fed into ducting centered over the hall and conducted air toward the extremity of the hall to an Energy Recovery Ventilator (ERV) in a mechanical room. Incoming outdoor ventilation passed across the hall-designated ERV located in a mechanical room. This incoming air crossed a preconditioning unit followed by heating and cooling coils before passing down the hall to be distributed by vents located centrally along the hall (10.2 cm ×10.2 cm) and above the door of each room (29.2 cm × 8.9 cm). The duct handling the incoming outdoor ventilation contained three 15-watt germicidal RCI ultraviolet (UV) Hg cells covering up to 189 m^2^. This HVAC design was similar in all halls of the building during normal operations.

**Figure 1:**
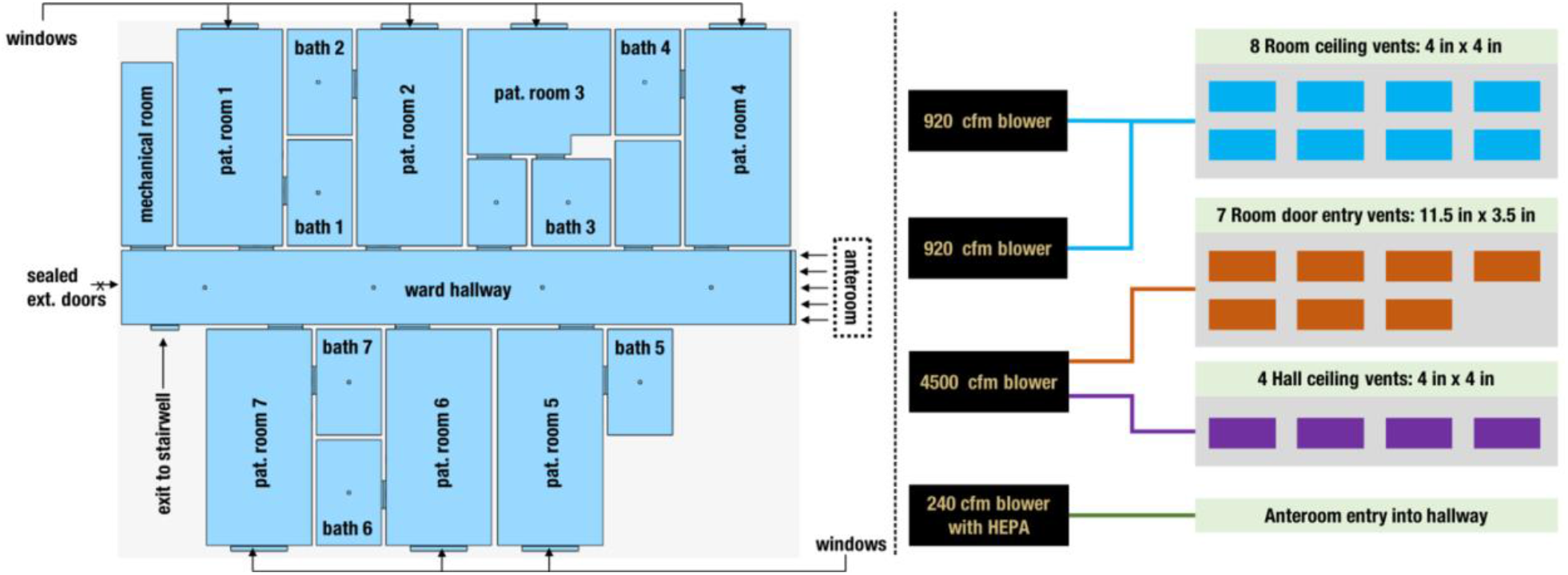
An illustration of the isolation space design. Right panel shows the floor plan layout of the isolation space (top view), with all patient rooms and entryways clearly identified. Vent locations are identified using square markers in appropriate locations. Left panel outlines the modified air handling system operating at 100% exhaust mode, showing how each blower connects to the vents in the isolation space.

### HVAC Modifications to Create Isolation Space

The HVAC and plastic barrier modifications were begun on April 2 and completed four days later on April 6. An overview of the modified air handling operations is presented in Figure 1 (right panel). Modifications were made to enhance the normal exhaust pattern by adding two new 0.43 m^3^/s (920 CFM) blowers within the ERV. Incoming outdoor ventilation fan was turned off using a breaker. A 0.51-m 2.12 m^3^/s (4500 CFM) exhaust fan was mounted to the outside of the intake grate on the east side of the building traditionally reserved for incoming outdoor ventilation, thus reversing the flow of air and converting the hall to 100% exhaust. To increase the exhausting of air within the hall, the preconditioning unit of the ERV was removed and the fire damper, located above the fire door, was closed within the ventilation system. Originally, HEPA filters were placed in the ERV, but due to restricted air flow these units proved prohibitive. The solution was to use Minimum Efficiency Reporting 8 filters and cordon off a 15.2-m radius around the exhaust vent on the east side. A long-term solution to purchase a germicidal UV light to be fitted into the ERV or mount HEPA filters to the exhausting air flows had yet to be completed.

### Anteroom Design

Located at the front of the hall and across the double fire doors, two sets of 3-sheet plastic barriers were installed constructing a temporary anteroom. The anteroom measured 2.4 m × 4.5 m in length. The fire door remained operational through the duration of this study. The barriers consisted of three overlapping sections of 152-micron plastic measuring 1.5 m × 2.5 m attached to the wall and ceiling using wood lathe. The barriers were rolled at the bottom just above carpet level and taped at the sides to identify separations for HCP access. The anteroom contained a 240 clean-air-delivery rate high-efficiency-particulate-air purifier fitted with 0.1-m ductwork to exhaust air into the hallway and toward the extremity of the wing (see for ref. Figure 1, left panel). A cascade of negative pressure zones with respect to both the isolation space (treatment hall), anteroom and the rest of the SNF was thus created.

Housekeeping staff was isolated from the hall with deliveries of linens being made to the anteroom. All housekeeping materials (i.e. vacuum, cleaning materials) were designated to the isolation wing and were not transported outside of negative isolation space. These items were stored within a laundering room located within the negative isolation space. The nursing staff was responsible for housekeeping tasks. A large waste collection container located outside the stairwell exit serviced the isolation wing so materials would not be transported through the building. Waste and laundry were both double bagged. Laundry was double bagged in dissolvable bags and removed from the anteroom daily. Donning and doffing of nursing personal protective equipment was done just outside the anteroom on the isolation side due to the need to reuse and clean PPE thus keeping the anteroom clean space. Pressure measurement equipment was located in an attached mechanical room within and adjacent to the anteroom.

During the initial testing of the system, winds were clocked by the onsite weather station at 33.8 kph. This data was supported by a national weather service automated observing station at a local airport. These winds had a significant effect on the exhaust fans and the overall ability to maintain negative pressure. Because of the capacity of this wind to generate a compromising positive pressure within the hall, a stairwell was selected for all resident entrances and exits from the wing. The design of the stairwell allowed the use of two separate doors, one to the outside and one to the hall in an air-lock type arrangement. This stairwell later served as an anteroom for donning and doffing PPE for visitors prior to entering the hall upon admission. Thus, patients and visitors could be admitted through the stairwell rear entrance to the negative pressure space. This designated entrance decreased the possible SARS-CoV-2 exposure of the broader facility.

### Pressure Measurements

Continuous measurement of pressure differentials was conducted through dates 05/14-05/21 using an Energy Conservatory (TEC) DG-700 dual-channel differential pressure instrument. Primary pressure differential measurements were taken: (a) between the anteroom interior and a nursing station located outside the anteroom away from the isolation space; and (b) between a location inside the isolation space close to anteroom entry and the anteroom interior. A set of secondary pressure differential measurements were included in the study in form of periodic spot-checks, where pressure differential was recorded between the anteroom and a location outside the building. The measurements during the study period were supplemented with an additional set of measurements spanning a 72-hour period during 05/23-05/25, where pressure differential was recorded between the nursing station outside the isolation space, and outside the building.

### Computational Fluid Dynamics Modeling

A computational fluid dynamics model was set up for evaluating air flow and negative pressure generation based on the HVAC modifications. The model was based on the finite volume method, using the cloud-based software suite SimScale (https://www.simscale.com). Full scale computer solid model of the entire isolation space ward was created using the web based CAD package OnShape (https://www.onshape.com), which was further discretized into a hex-dominant numerical grid. Based on built in SimScale modules, a steady compressible flow solver was set-up using the k-omega model for turbulence, to model the flow of air throughout the isolation space. Air handling vents in the patient rooms and along the hallway were modeled using velocity outlet boundary conditions. All vent boundary conditions were derived using reduced order volumetric flow distribution calculations (*see supplementary material*). Narrow slits were included at the anteroom entry, and the stairwell doors opposite the mechanical room, which were also modeled as pressure inlet boundary conditions. Lastly, a set of pressure inflow boundary conditions were assigned to each patient room window, modelled also as narrow slits, to account for infiltration airflow. Resultant fluid flow velocity and pressure field data were then used to drive a Lagrangian particle-based model for simulating viral particle dynamics within the isolation ward. A fixed number concentration of viral particles was released at various locations in the isolation space, and their trajectories and final locations over a 4-5-minute window were computed. Additional details regarding aspects of the computational methodology are included in supplementary materials.

## Results

### Measured Pressure Differentials Showed Sustained Negative Pressure

The measured pressure differentials between: (a) anteroom and outside the isolation space; as well as (b) between inside the isolation space and the anteroom; reveal sustained continuous negative pressures into the isolation space across the anteroom throughout the duration of the study. Hourly median values for pressure differentials (a) an (b) above centered around -1.27 Pa (std. dev.: 0.1 Pa) and -1.05 Pa (std. dev.: 0.16 Pa) respectively. Figure 2 illustrates the measured pressure differential across the anteroom entry into the isolation space. The bottom panel represents an hourly max-min envelope for one of the observation dates (May 21). Specifically, the median values of all observations within one hour are represented using the solid line, while the shaded region presents the upper and lower bounds. Median hourly pressure differentials for each observation date for both (a) and (b) above are presented on Figure 2 top panels. These exclude the spot-check measurements of pressure differentials as described in the Methods. All median pressure differentials are negative, implying the isolation space is at a lower pressure than the anteroom, thereby preventing any air flow out from the isolation space towards the anteroom and rest of the facility. Sporadic positive pressure differentials were observed during the study, which explains the upper bound in Figure 2, bottom panel, being positive for certain hours during the day. This is likely due to movement of healthcare workers and staff across the anteroom and opening/closing of anteroom entry for access into the isolation space. These measurements are further supported by on-site observations that the anteroom plastic sheet entryway remained continuously curved inwards into the isolation space, indicating pressure on the anteroom side to be higher than on the isolation space side.

**Figure 2:**
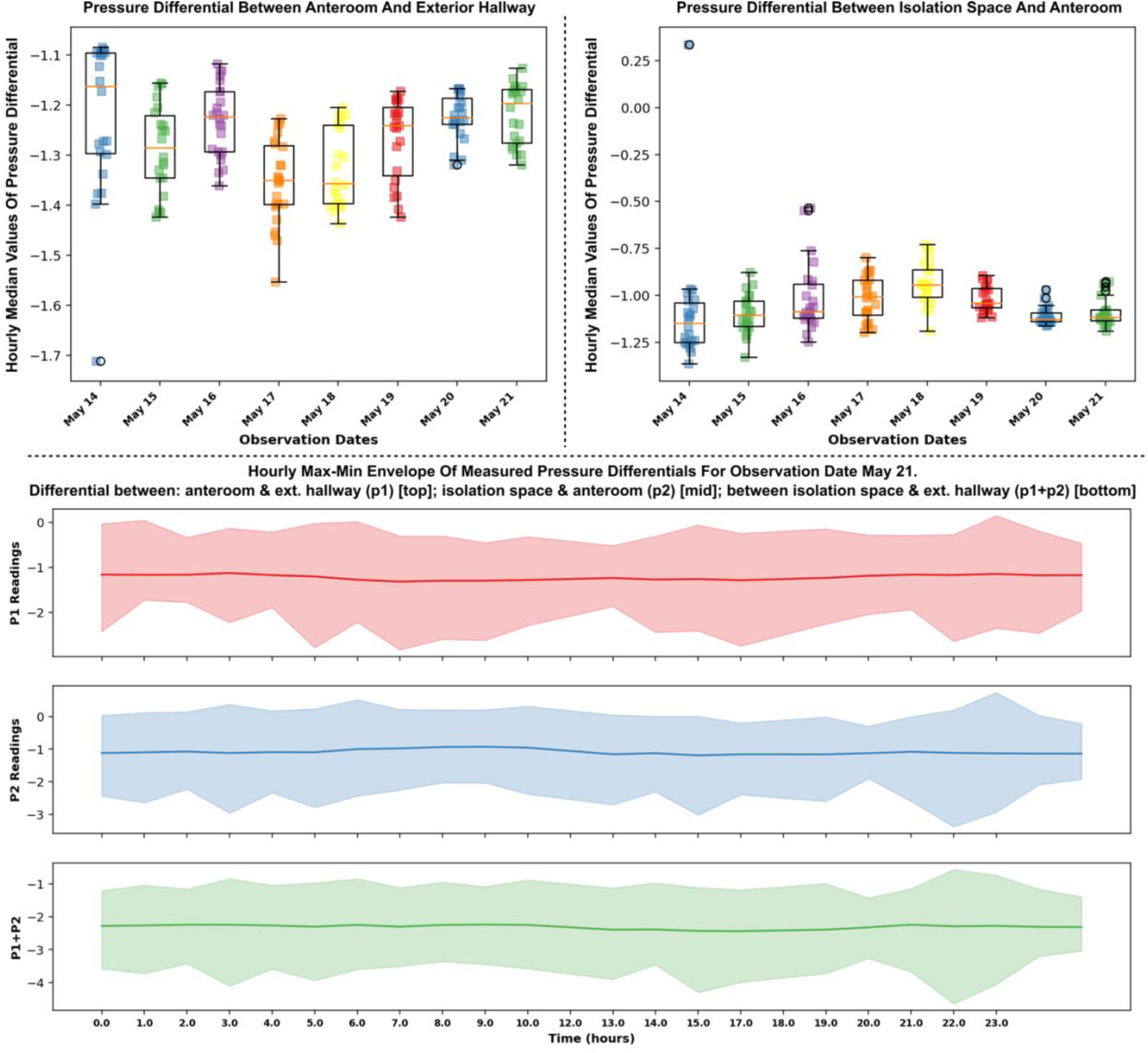
Measured pressure differential data during observation period May 14 – May 21. Top left panel shows hourly median pressure differential between anteroom and exterior hallway, while top right panel shows the same for pressure differential between isolation space and anteroom. Bottom panel shows the measured pressure differentials, represented using a max-min envelope, for one of the observation dates (May 21) to illustrate the hourly variations. Data for remaining observation dates presented in supplementary information.

### Computational Model Validated Measured Negative Pressure

A pressure differential was computed from the CFD simulation data with reference to the pressure at the anteroom entry, based on measured pressure data across the isolation space. Spatial variation of this computed pressure differential throughout the isolation space is visualized in Figure 3, along a slice taken midway through the height of the isolation space, viewed from the top. The data presented here assumed that all patient room doors were open, but the anteroom entryway was operational (as described earlier), and the stairwell doors at the end of the hallway as well as all windows were closed. Values of the computed pressure differential are also presented in Figure 3 (bottom) along a line spanning the entire length of the isolation space hallway (colored based on flow velocity magnitudes). Since the pressure measurement probe inside the isolation space was placed close to the anteroom entry, we used the obtained pressure differential values close to the anteroom entry for cross-validation with measured data. The computed pressure differential values show excellent agreement with measured pressure differential values as shown in Figure 2 (top right panel) in form of distribution of hourly median pressure differential measurements for each of the observation dates. It is also observed from Figure 3 that the pressure distribution has significant variations spatially across the isolation space. However, so long as the pressure differential across the anteroom entry into the isolation space remains negative, the airflow remains unidirectional into (and not out of) the isolation space, creating effective containment.

**Figure 3:**
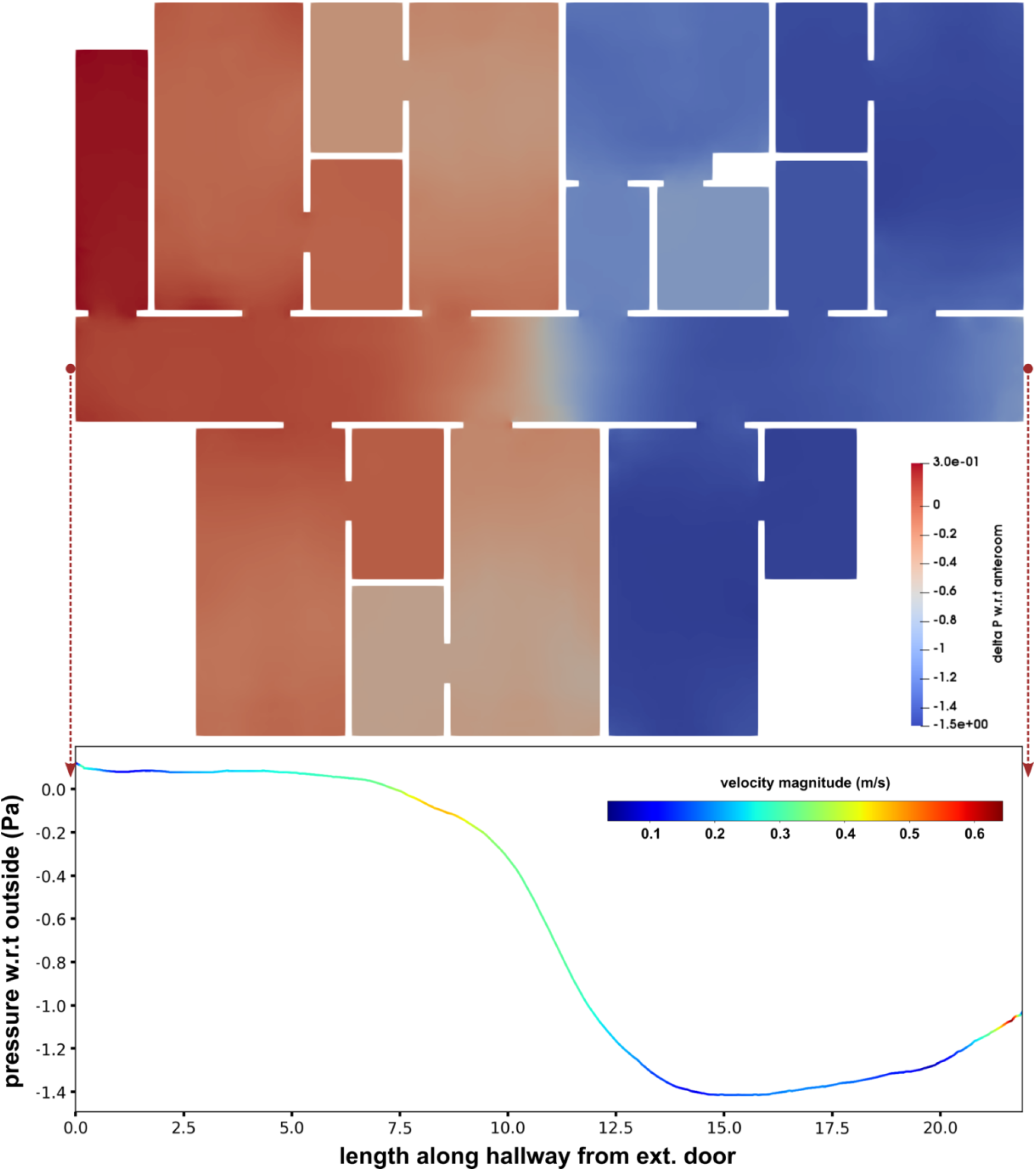
Pressure differential obtained from computational modeling, visualized across the isolation space. The top panel shows the pressure differential along the mid-section of the isolation space viewed from the top. Bottom panel presents the differential along the middle of the hallway on the mid-section, colored using the flow velocity magnitudes. Pressure differential close to anteroom entry is -1.0 - -1.1 Pa which matches the measured data shown in Figure 2 (top right).

### Modeling Further Elucidated Air Flow and Viral Particle Transport Patterns

Results from computational modeling provided additional insights into the air flow pattern within the isolation space and indicated possible modes of viral particle transport across the rooms and hallway. Figure 4 presents the flow velocity magnitudes and local circulation patterns across the slice taken midway through the height of the isolation space (same as in Figure 3), for the scenario where all room doors are open. The flow pattern is visualized using line integration convolution (LIC), which concurrently illustrates flow magnitudes as well as local vortex cores. Slow flow velocities in the order of 0.1-0.5 m/s span the entire hallway with local circulatory flow regions influenced by the layout of the isolation space, the operation of air handling and HVAC systems, and infiltration from the windows. Figure 5 shows results from two sets of numerical experiments on viral particle transport. For the first experiment (Case A), particles are released in three of the patient rooms, and for the second experiment (Case B), particles are released at two locations along the hallway. With all patient room doors kept open, these experiments show the extent to which released particles can spread in the isolation space - from one room into another, as well as from the rooms into the hallway and vice versa. Results shown in Figure 5 illustrate that when patient room doors are open, viral particles may spread from room to room, as well as into the hallway – possibly affecting other patients and healthcare workers. Additionally, viral particles shed in the hallway may infiltrate into patient rooms. These observations indicate that despite establishment of isolation space via sustained negative pressures, careful observation of protective measures within the isolation space (as conducted in this study) remains critical to prevent viral transmission amongst healthcare workers. We note that here we have not accounted for detailed and coupled flow through the ductways. Consequently, any particle that exits from the vent into the ductway are considered to be ‘removed’ from the simulation. In future studies we intend to resolve this further to improve accuracy of viral particle transmission estimates.

**Figure 4:**
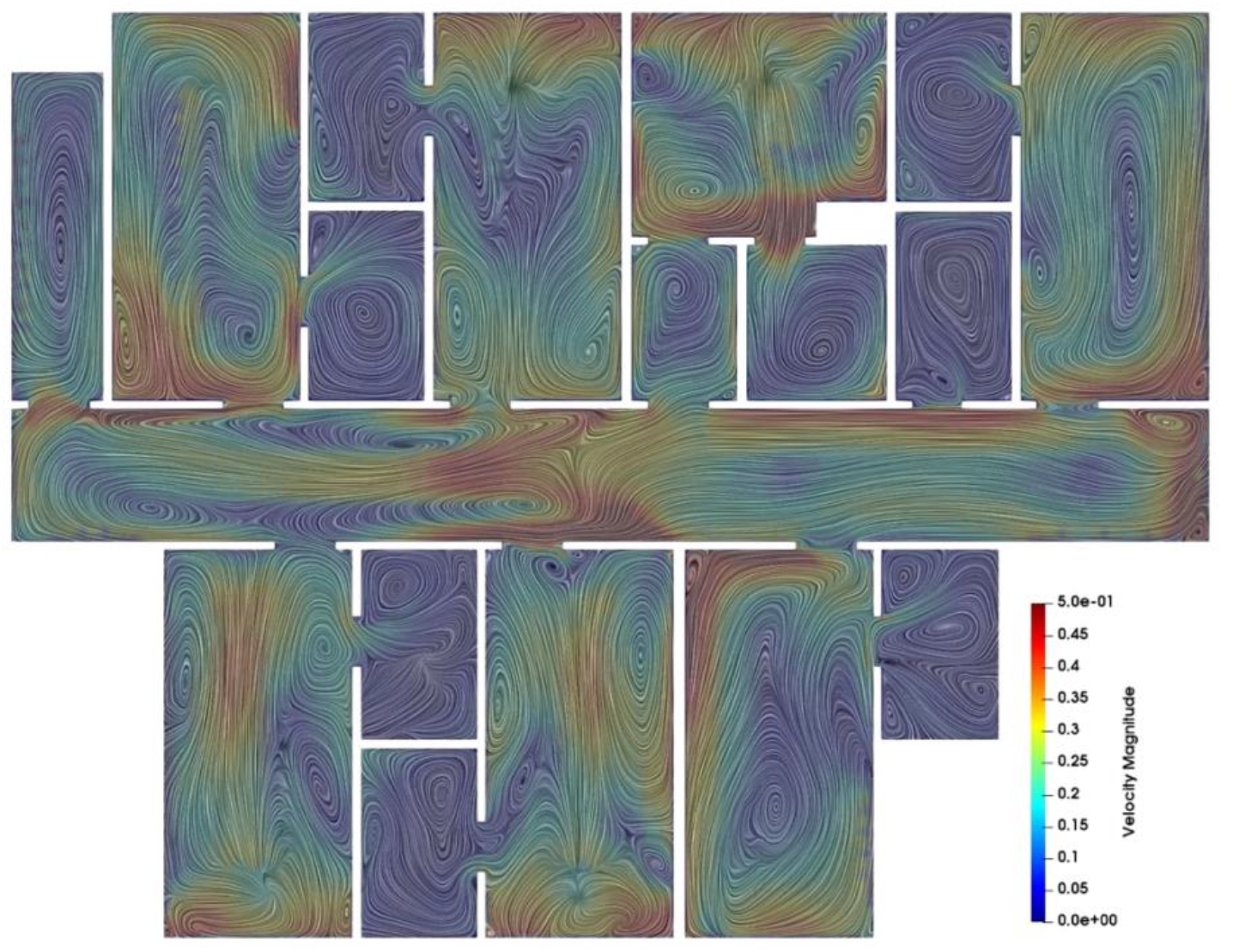
Illustration of spatially varying air flow patterns inside the isolation space, viewed using line integration convolution (LIC), which clearly shows flow velocity magnitudes (in the range 0.1-0.5 m/s) as well as local circulation patterns. Flow is visualized along the same mid-section as in Figure 3.

**Figure 5:**
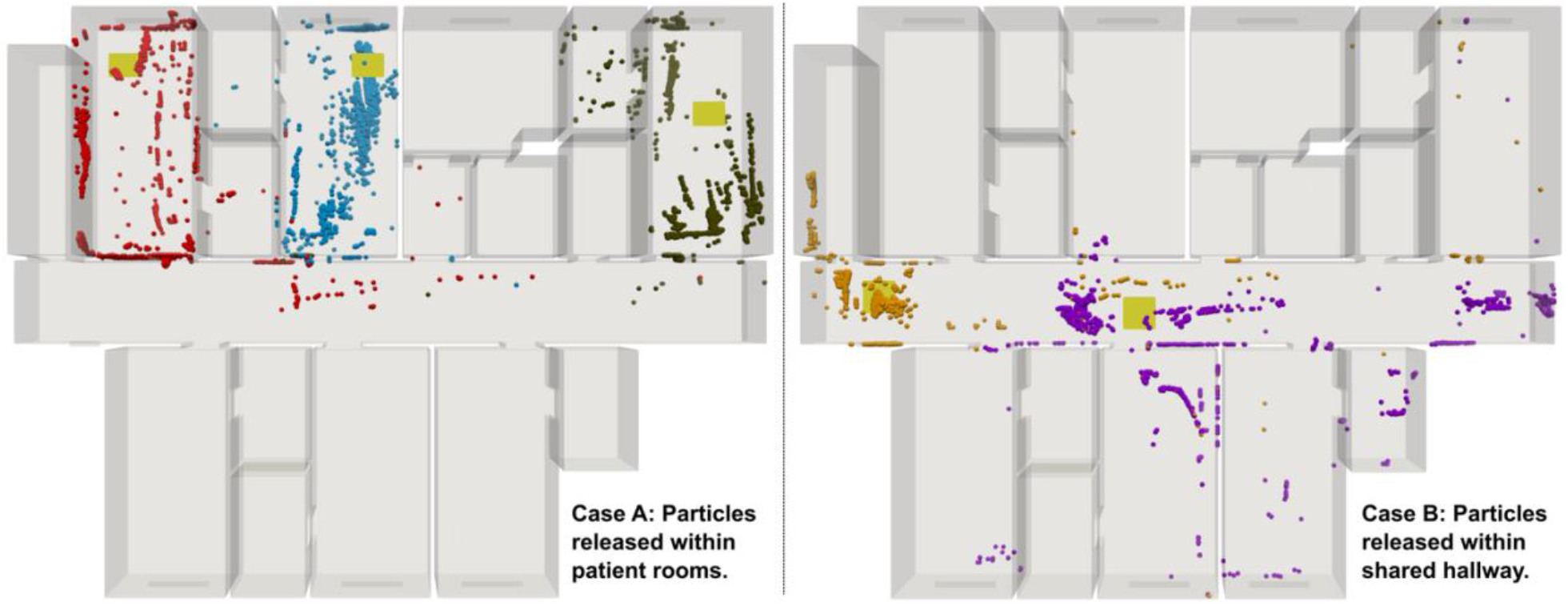
Results from Lagrangian particle simulations for viral particle movement in the isolation space. Case A (left panel) shows scenarios where particles are released within patient rooms. Case B (right panel) shows scenarios where particles are released in the hallway. Release locations are marked using yellow patches.

### Isolation Space Was Successful in Preventing Any Form of Transmission

The negative pressure isolation space was established April 6, 2020. Experimentation and data collection began May 14, 2020. Census data during our study showed between 2-4 known SARS-CoV-2 patients resided within the isolation space for the duration of the study. A designated team of healthcare workers were assigned to task the negative pressure space. The workers consisted of nursing staff (RNs, LPNs, CNAs), physical therapy staff, a respiratory therapist, and a nurse practitioner. While some of the nursing staff exclusively worked in the isolation space, they did have significant contact at shared nursing stations outside of the space with those working off the unit. The therapy staff and nurse practitioner worked throughout the facility. The nurse practitioner conducted the SARS-CoV-2 testing. Staff entering the negative pressure space were provided PPE training in donning and doffing directed by the respiratory therapist and completed over two days. Goggles, N95 masks, and gowns were issued to staff entering the isolation space. Nursing care of these residents ranged from vital signs q4h through asymptomatic. Over the study admissions and discharges occurred. The facility received its first known positive COVID-19 patient as a hospital transfer on April 8. From that date onward, and over the course of this study, the facility acquired no known positive results within its own population. None of the healthcare workers assigned to work the isolation space while treating or in proximity to COVID-19 patients presented with symptoms or tested positive thereafter.

According to the Pennsylvania Department of Health, as of June 23, 2020, of the 80,810 reported cases in Pennsylvania 17,394 resident cases and 3,103 worker cases were reported with 4467 deaths in 671 facilities. Facilities consisted of both personal care and long-term care communities. The total number of cases in Lancaster County on June 23 was 988 resident and 286 worker cases with 251 deaths (2020, June 23). As of June 23, the isolation space has consistently maintained a population of known COVID-19 residents ranging from between 2 and 6 individuals. Residents are routinely treated and discharged from the isolation space after negative test results are acquired. As of June 23, 14 confirmed (PCR testing) SARS-CoV-2 residents had been treated in the negative isolation space established in this study, and the facility had utilized the isolation space for a total of 21 individuals. These additional individuals had been quarantined prior to the widespread availability and access to testing, in the isolation space as suspected cases. At the time, these individuals represented a possible transmission to the facility due to external exposure at another facility such as a hospital. None of the quarantined individuals transferred from the isolation space, with stays lasting between 2-14 days, later tested positive or presented symptoms. One individual that was suspected and placed in isolation space quarantine tested positive on day 10. According to the PA DOH this individual was not considered as facility onset. The PA-HAN-509 Facility-onset SARS-CoV-2 does not include infections in residents with known exposure outside of the nursing home or those that become positive within 14 days after admission, when placed into Transmission-Based Precautions. The placement of this individual into transmission-based precautions, which in this study included the negative isolation space, represented a significant role the isolation space played in mitigating possible transmission of the virus within the facility.

## Discussion

Skilled Nursing Facilities offer available surge capacity under pandemic scenarios. Elderly patients require significant and disproportionate health services within a regional health network (Institute of Medicine, 2008). In addition, treating nursing home residents in place offers considerable advantages due to the risk/benefit decisions surrounding transfer and admission to emergency departments (EDs). The transition of older adults from a nursing home setting to the hospital involves significant challenges surrounding structured communication between nursing homes and EDs. These issues involve, but are not limited to, cognitive impairment, missing documentation involving baseline cognition, and goals of care (Levine, 2020; Hustey, 2010). The implementation of a negative pressure isolation space within a SNF alleviates hospital admissions and staffing loads, thus generating surge capacity without jeopardizing healthcare standards associated with transfers. Previous studies in nursing homes during outbreaks have identified lower culture confirmed influenza A (H3N2) cases where unique ventilation and increased space per resident exists (Drinka 1996).

Nursing home providers often have access to knowledgeable professionals including respiratory therapists available to direct and train staff in effective PPE procedures. Beyond the availability of skilled nursing staff familiar with resident behaviors and treatments, many existing SNFs are architecturally designed according to hospital models with rooms branching off central, hospital-like corridors with corresponding HVAC systems. As such, we predict that an in-house modification to an established HVAC system will be able to sustain negative pressures that meet or exceed CDC guidelines as previously reported in hospital modifications (Miller et al. 2017). Modifications to existing infrastructure can typically be less resource-intensive and can be rapidly conducted with existing expertise and promptly utilized. Full-scale computational modeling can be used to evaluate efficacy of such modifications, and validate sustained negative pressures, as demonstrated here.

Within the negative pressure area that we designed the staff will need to wear PPE, due to the possibility of positive pressures within rooms and hallways. Additional design and testing is needed to address this concern and strategies include increasing exhaust airflow from the bathrooms to maintain a negative pressure within the bathroom and patient room, install upper-room germicidal ultraviolet lights within the area to inactivate any airborne virus, and use of air purifiers to exhaust air from each room to the outdoors.

## Conclusions

We implemented a negative pressure isolation space within a SNF to contain SARS-CoV-2 transmission, by modifying existing HVAC systems. Our modifications were able to achieve and sustain negative pressures generating an ideal isolation unit for the residents with confirmed COVID-19 disease within the nursing facility. Consistency between both on-site measurements and modeling simulations confirm the efficacy of planned modifications to the HVAC system to sustain negative pressures that meet or exceed CDC guidelines. Furthermore, the community benefited from the expansion of available health services and decreased community spread in facilities that lacked sufficient mitigation strategies. Residents avoided the stress associated with transfers to and from emergency facilities. Thus, the resulting isolation space operated successfully through the ongoing pandemic.

## Data Availability

Data is available upon request.

## Supporting Information

Supplementary information on details regarding the computational modeling approach and supplementary pressure measurements data has been included in an additional document: *supplementary-modeling-data*.*pdf*. In addition, a document describing staffing procedures for the facility as per DOH, CMS, and CDC recommendations, has been included in document: *staffing-prcedure*.*pdf*

## Acknowledgments

We thank the Administration and Facilities management at Fairmount Homes, John Becker and Jerry Lile for their significant support in helping to engineer and support the project along with Therapy Trip LLC and Goodville Mutual Casualty Company for the initial funding to spur the project. Jen Roberts Don and Susan Noriega Fairmount Homes for communication and access to nursing information and staff. Daniel Mergner RCT for help with preliminary data collection and Zachary Zimmerman for thoughtful discussions about language and communication. We thank SimScale for help with license, and students Akshita Sahni and Chayut Teeraratkul for their assistance during initial evaluation of SimScale for this study.

## Author Contributions

Conceived and designed the experiments: CDS, SLM, NSC. Performed the CAD modeling: JW. Performed CFD and particle simulations: JW, DM. Developed pressure measurement workflow: NSC. Analyzed the data: DM, SLM. Wrote the paper: CDS, DM, SLM.

